# DEVELOPMENT OF AN IDENTIFICATION METHOD FOR *Leishmania* spp BASED ON REAL-TIME PCR WITH HIGH-RESOLUTION MELTING

**DOI:** 10.1101/2024.08.30.24312710

**Authors:** Rafael Villareal Julio, Brayan E. Ordoñez, Gabriel A. Pérez, Valeria Manjarrez, Katia Villarreal Julio, Carlos Muskuz

**Affiliations:** PECET, Faculty of Medicine of the University of Antioquia, Medellin, Colombia; BIOTECH MOLECULAR, Molecular, Genetic and Computational Biology Unit, Medellín, Colombia; University of Antioquia, School of Medicine, undergraduate medicine, Medellín, Colombia; Cooperative University of Colombia, undergraduate medicine, Medellín, Colombia

**Keywords:** *Leishmania*, diagnosis, identification, real-time PCR, melting curves

## Abstract

**Introduction:** *Leishmaniasis* is a disease caused by parasites of the genus *Leishmania*. In Colombia, 6 pathogenic species have been reported. Traditional parasitological diagnosis based on the observation of parasites does not allow the species to be identified, which is why biochemical or molecular methods must be used, including conventional PCR, but this has some limitations and requires long periods of time to obtain results., which are sometimes inconclusive.

**General Objective:** Implement a methodology based on PCR-HRM, which allows the simultaneous detection and identification of parasites of the *Leishmania* genus in clinical samples from patients or parasites in cultures.

**Materials and methods:** A PCR-HRM was standardized, through which 237 clinical samples were evaluated, 98 classified as positive and 139 as parasitologically negative by direct and/or culture. The typings were compared with a previously standardized PCR-RFLP.

**Results:** It was possible to implement a PCR-HRM for the diagnosis and identification of *Leishmania* species, obtaining 100% agreement with the typings obtained by PCR-RFLP. It was even possible to detect and identify the parasite in samples diagnosed as negative by conventional methods. It was found that, with a reliability percentage greater than 95%, 91 samples out of 98 were typed, of which 81.63% of the cases were L. panamensis, 11.22% were L. braziliensis and 7.14 were indeterminate.% of cases.

**Conclusions:** PCR-HRM is a good method that allows the identification of the most prevalent species in Colombia, comparing specific mean denaturation temperatures according to the *Leishmania* species involved.

## 1. Introduction

*Leishmaniasis* is a disease caused by protozoa of the genus *Leishmania*, within which at least 15 species are pathogenic to humans.(1). The infection is endemic in 98 countries located in tropical and subtropical regions (2). Three basic clinical forms of the infection have been described: Cutaneous, mucosal and visceral *Leishmaniasis*, the latter being the most severe form of the disease (3).

A timely diagnosis and, if possible, accompanied by the identification of the species is necessary for the administration of adequate treatment and follow-up. However, traditional diagnostic methods such as direct examination and culture generally lack adequate diagnostic sensitivity and do not allow identification of the species. Traditional diagnostic methods have been replaced by molecular techniques, especially PCR-based methods, since it is a rapid, sensitive and specific technique.(4), as a diagnostic method, but to identify the species a post-PCR evaluation such as RFLP (restriction fragment size polymorphism) is normally required (5), hybridization(6)or sequencing (7).

Although these advances in molecular techniques show better results compared to conventional methodologies, they are still somewhat costly if species identification needs to be carried out in addition to diagnosis. That is why the need has arisen to use even variations of current methodologies, which allow not only the diagnosis but also the identification of the species to be carried out more quickly and, in the case of identification, more reliably. It is known that real-time PCR, which under optimal conditions presents greater sensitivity and high specificity than conventional PCRs (3), when coupled to high-resolution mean denaturation temperature curves (HRM-High Resolution Melting) is useful, in our case, for the discrimination of species either from cultures of the microorganism or directly from the clinical sample, which is advantageous for microorganisms that are not isolated or even microorganisms that can be isolated but that For some reasons, it is not possible to cultivate it for the subsequent identification of the species. HRM curves measure the rate of dissociation from double-stranded DNA to single-stranded DNA with increases in temperature and recording changes in fluorescence corresponding to fragment size, GC content, complementarity, and thermodynamics of the closest nucleotides.(8).

Taking into account the need to implement a methodology that allows diagnosis and identification of species in a single step and based on the sensitivity, specificity, speed of PCR in real time and the possibility of identifying variations in a sequence using temperature curves of high resolution medium denaturation, the present work aimed to implement a qPCR-HRM that allows diagnosing and identifying the infecting species in clinical samples from patients (9).

## 2. Materials and methods

### Study population

Patients treated at the PECET office of the University of Antioquia and coming from endemic areas of Colombia or who visited endemic areas in the last 6 months and with the presence of lesions compatible with cutaneous *Leishmaniasis*, not receiving treatment at the time of consultation, were included. and those who expressed their voluntary participation in the study, by signing an informed consent, those who did not meet the aforementioned inclusion criteria were excluded. The sample size was calculated using Epidat V 3.1, according to the QUADAS fundamentals, and the ratio of non-sick/sick patients was 10/7 in order to obtain results with a sensitivity greater than or equal to 95%, specificity greater than or equal to at 90% and with a confidence level of 95%. The study is prospective and the samples were processed blindly.

### Sampling

The samples were collected between the years 2012 and 2014. All samples were processed at the PECET laboratory facilities. The processing of samples by the different diagnostic and typing methodologies was carried out by experts, with extensive experience in each of the methodologies, each process was carried out separately, and each person was in charge of only one methodology. The following types of samples were taken from each patient on the same day of the consultation and simultaneously: an aspirate for culture, a scraping for direct examination, which is the Gold standard in the disease, and a scraping for DNA extraction and PCR (Conventional and qPCR-HRM) which were performed in triplicate.

For direct examination, the scraping sample was fixed with methanol and subsequently stained with Giemsa for 15 minutes and washed with water and read by two diagnostic experts independently.(1). For culture, an aspirate is taken from the lesion using a tuberculin syringe containing a liquid phase, and this is cultured in NNN medium at 26°C for at least 30 days, evaluating the presence of parasites by microscopy every week and passing the samples. fresh NNN medium cultures(1).

For PCRs, DNA extraction was performed using DNeasy® Blood & Tissue (Qiagen, Hilden, Germany) following the protocol established by the manufacturer. The DNA obtained was quantified in a Nanodrop (ND1000, Thermo Scientific, Wilmington, USA). The quality of the purified DNA was further evaluated by electrophoresis in a 1% agarose gel containing 0.5 µg/ml ethidium bromide and visualized on an image documenter.(10).

### Target sequence selection and oligonucleotide design for PCR-HRM

For real-time PCR coupled to HRM, oligonucleotides were designed that amplify a previously selected region based on multiple alignments using the sequences reported in the NCBI and Tritryp. The alignments were carried out with T-coffee and viewing in MEGA7. Using these alignments, differences were searched and evidenced in the nucleotide sequence of the DNA polymerase 2 gene between the 6 different species of *Leishmania* spp present in Colombia. The oligonucleotides were designed using the Primer3Plus program (http://www.bioinformatics.nl/cgi-bin/primer3plus/primer3plus.cgi) and the quality of the oligonucleotide design was evaluated by oligoanalyzer (https://www.idtDNA.com/calc/analyzer), to determine the non-formation of homo- and heterodimers in addition to hairpins. The specificity of the oligonucleotides was analyzed by Blastn.

### Real-time PCR and real-time PCR with high-resolution melting curve analysis

HRM-PCR was performed using 1 µl of each oligonucleotide (10 µM), 12.5 µl of 2X Master Mix (HRM PCR Qiagen, Hilden, Germany), 9.5 µl of DNase-free water (Qiagen, Hilden, Germany) and 1 µl of DNA (10-20 ng/µl) for a final volume of 25 µl per sample. The reaction was carried out in the Rotor-Gene 6000 Corbett Life Science thermocycler (Qiagen, Hilden, Germany). The thermal profile used was an initial denaturation cycle at 95°C for 5 minutes, followed by 40 cycles of denaturation at 95°C for 10 seconds, alignment at 57°C for 40 seconds and extension at 72°C for 10 seconds.. The analysis of high resolution denaturation mean temperatures (HRM) was carried out between 78ºC and 90ºC, increasing 0.1°C every 2 seconds to achieve denaturation of the amplicons, and taking fluorescence readings at each temperature increase. The analyzes were performed using the High Resolution Fusion Q software program (Qiagen, Hilden, Germany).

### Linearity, detection limit, reproducibility and analytical specificity

To verify the linearity of the PCR-HRM, a standard temperature curve was constructed from DNA isolated from *Leishmania* panamensis parasites. The curve was built from 390,625 to 5 parasites using base 5 dilutions.

To perform the detection limit analysis of conventional PCR and real-time PCR-HRM, 1:10 serial dilutions of L. panamensis DNA were used, starting from a concentration of 10 ng/µl until reaching a dilution of 1 :10,000,000 equivalent to a concentration of 1 fg/µl of parasite DNA.

The analytical specificity analysis was carried out using DNA from Trypanosoma cruzi (AC29), Sporothrix schenckii, Staphylococcus aureus and Mycobacterium tuberculosis and as a negative control DNA from healthy people not infected with *Leishmania* spp was used.

The reproducibility analysis was performed by processing all samples in triplicate and evaluating the agreement in both diagnosis and typing between the three replicates of each sample.

### Evaluation of human DNA interference

3 concentrations of DNA were taken from the parasites; 0.7 ng/µL, 3.5 ng/µL and 17.6 ng/µL corresponding to 3,125, 15,625 and 78,125 parasites, respectively. Additionally, DNA extracted from human mononuclear cells was added to each sample, up to 20 ng/µL of total DNA, and PCR-HRM was performed following the protocol described above.

### High resolution melting curve analysis (HRM)

It was carried out with the Rotor-Gene Q Series Software 2.0.2 (Build 4) program using the High Resolution Fusion Q software module (Qiagen, Hilden, Germany). In order to perform an optimal HRM analysis, it was checked that the amplification data met the following characteristics: a) all amplification curves had a Ct value < 30, b) all amplification curves had to reach a similar plateau phase, c) between samples, the Ct value should not vary more than 5 Ct units or five cycles, to guarantee that similar concentrations of DNA would exist in each sample. The mean denaturation temperature curves were analyzed with the Gene Scanning program.

Simultaneously with the running of all samples by PCR-HRM, all samples were also evaluated using a PCR for detection and PCR-RFLP for the Identification: Conventional PCR was performed using the Hsp70-N protocol described by Montalvo, *et al*.(5). 1 µl at 20 ng/µl was amplified as follows: Initial denaturation at 95ºC for 5 minutes, followed by 35 cycles of denaturation at 95ºC for 45 seconds, oligonucleotide annealing at 61ºC for 45 seconds and extension at 72ºC for 45 seconds, with a final extension at 72ºC for 8 minutes in a C1000 thermocycler (Bio-Rad, Foster City, CA, USA). The amplification product was visualized on a 1% agarose gel, where an amplicon of 593 bp should be observed. The enzymatic digestion for the RFLP was performed in a final volume of 10 µl, using 8.5 µl of the amplicon, 1 µl (1X) of Tango buffer and 0.5 µl (2.5 U) of the restriction enzyme BsaJI (MBI Ferments). The reaction mixture was incubated at 55°C for 3 hours. Analysis of the digestion of the PCR products was performed by electrophoresis in a 4% agarose gel (AMRESCO, Ohio, USA), which was run at 70 V for 2 hours.

### Statistical analysis

The sample size for validation of the diagnostic tests was calculated assuming a sensitivity of 95%, a specificity of 90%, the ratio of sick and non-ill patients of 1.43 and a confidence level of 95%, the sensitivity, specificity, values Positive predictive values were calculated using Epidat® 3.1.

Diagnostic agreement was calculated using a Kappa index between conventional PCR versus real-time PCR, real-time PCR versus the GOLD Standard (Direct + Culture).

## 3. Results

A sample size of 139 healthy people and 98 sick people was calculated, estimating a precision of 5%, for a total of 237 patients (Table 1.)

**Table 1.**
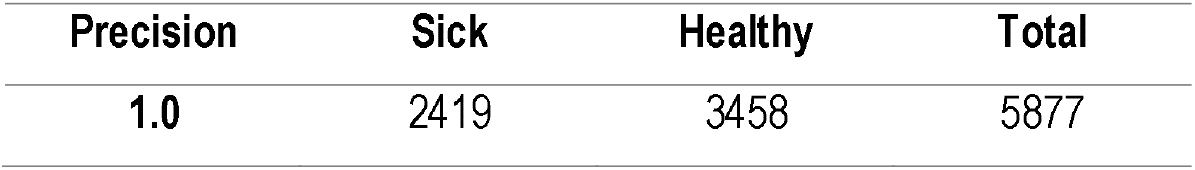
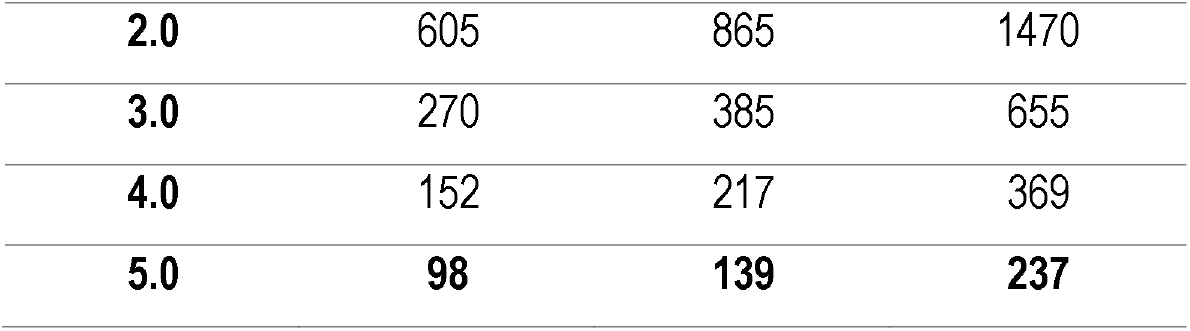
Estimation of sample size.

### Direct examination and culture

Of the initial 237 samples, 98 were positive by microscopy while only 48 samples were positive by culture (Table 2).

**Table 2.** Results of the different methods used for the diagnosis of cutaneous *Leishmaniasis* in clinical samples.

| **Result** | **direct**<br>**examination** | **Crop** | **Hsp70N**<br>**PCR** | **PCR-**<br>**HRM** |
| --- | --- | --- | --- | --- |
| Negative | 139 | 189 | 150 | 139 |
| Positive | 98 | 48 | 87 | 98 |
| **Total** | **237** | **237** | **237** | **237** |

#### DNA extraction

An average of 80 ng/µL of DNA was extracted in each sample and degradation was not observed in any of the samples.

#### Bioinformatic analysis

Using the sequences reported for the DNA polymerase 2 gene in: L. panamensis, L. braziliensis, L. guyanensis, L. amazonensis, L. mexicana and L. infantum taking as reference the DNA polymerase 2 gene with accession number AJ304944 of L. guyanensis it was determined that in the region between536and649 nucleotidesdifferences were found in the sequences in the composition of the bases between each species. To amplify this region where the variations were found, the oligonucleotides FW: AGGAGGATGGCAAGCGGAAG and RV: GCGACGGGTACAGGGAGTTG were designed. These bioinformatic analyzes were also carried out on other sequences in order to finally select for diagnosis and differentiation the sequence that yielded the best results experimentally (Figure 1).

**Figure 1.**
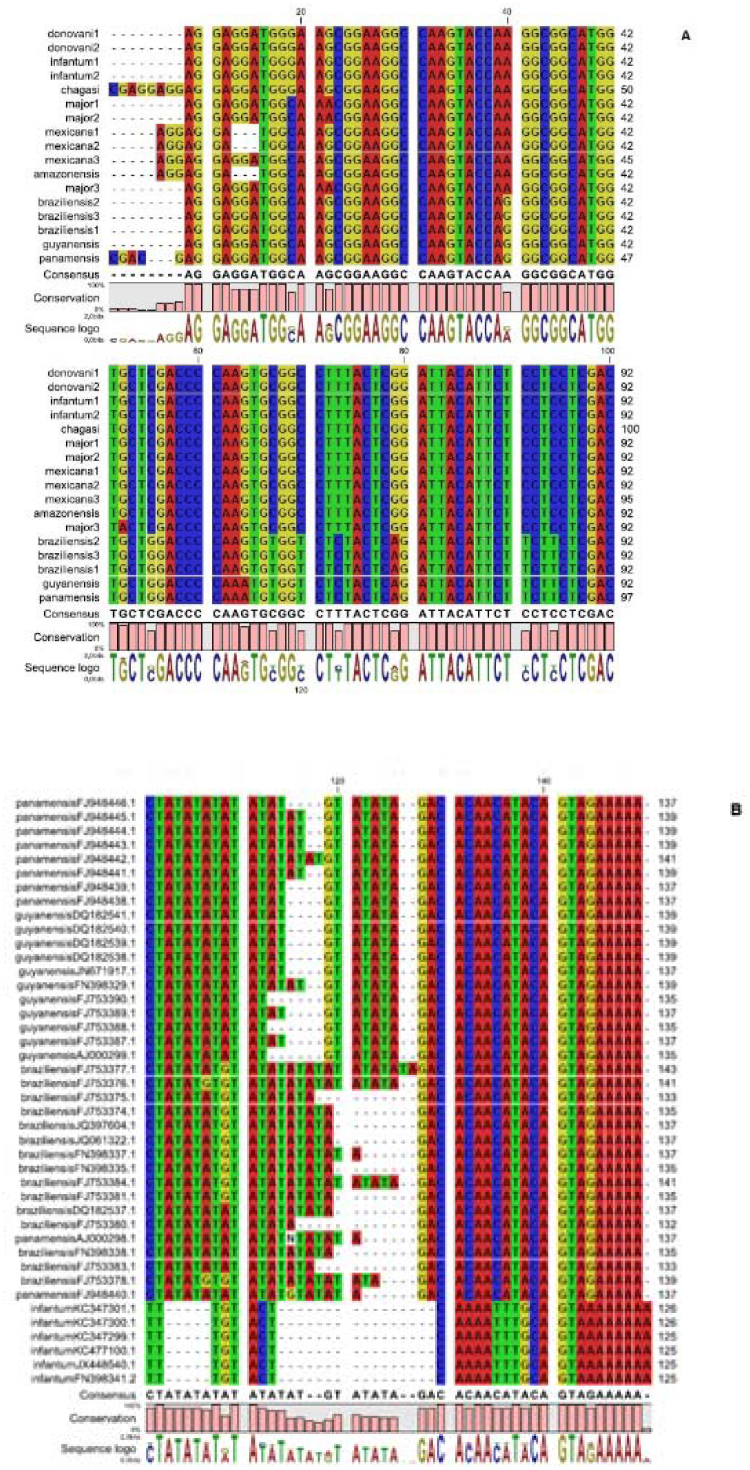
Multiple alignment of the sequences of the ADNpol 2 gene from different *Leishmania* species and visualized using Jalview. Panel A. Alignment of the conserved regions within and between species in the DNA polymerase 2 gene. Panel B. The alignment shows greater variability in the regions than those seen inthe ITS1 region. Conserved regions within and between species are observed in the sequence of the ITS 1 region. Changes in color indicate polymorphisms according to the base that changes (Adenine: red, Cytocine: blue, Guanine: yellow and Thymine: green) and the Blank spaces correspond to deletions in some of the sequences or insertions in the others.

#### Diagnosis and identification by PCR-RFLP

By amplifying the N-terminal region of the HSP70 gene (HSP70N), a total of 87 samples of the 237 were diagnosed as positive. Of these 71 samples were identified as L. panamensis (81.6%), 2 samples as L. braziliensis (2.29%) and 14 (16.09%) samples were classified as indeterminate, because the RFLP patterns were not compatible with any of them. reference standards.

#### Real-time PCR and high resolution mean denaturation temperature analysis-PCR-HRM

Prior to analyzing the performance of the test in clinical samples, the analytical specificity, analytical sensitivity, linearity of the method, and human DNA interference were evaluated. Initially using DNA from the 6 species of *Leishmania* that have been reported in Colombia as a control, the expected fragment was amplified and not amplified in DNA from Sporothrix schenckii, Staphylococcus aureus and Mycobacterium tuberculosis. It was possible to detect from 1 fg of parasite DNA, while by conventional PCR it was only possible to amplify from 1000 fg. Using 20 ng of DNA, in a parasite DNA/human DNA ratio as follows: 5/1, 1/5 and 0.1/5, it was possible to amplify the expected fragment, without affecting the specificity.

**Image 1.**
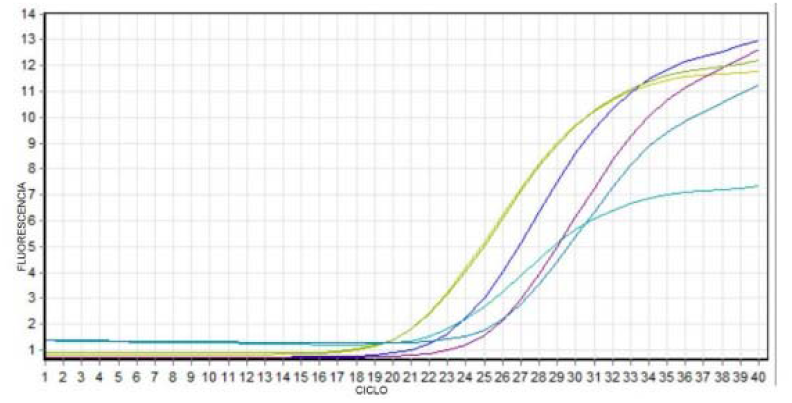
Determination of human DNA interference in the detection of parasitic DNA. The comparison of 3 different concentrations of DNA from*L. panamensis*:17.6 ng/μL 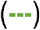, 3.5 ng/μL 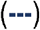 and 0.7 ng/μL 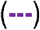 vs 17.6 ng/μL + 2.4 ng/μL human DNA 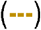, 3.5 ng/μL + 16.5 ng/μL human DNA 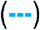 and 0.7 ng/μL + 19.3 ng/μL of human DNA 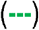.

#### High resolution mean (HRM) denaturation temperature curve analysis

Taking into account that in Colombia 98% of the cases correspond to L. panamensis, L. braziliensis and L. guyanensis, the differentiation of these three species was validated using PCR-HRM (Image 2.), achieving typing with the established protocol. 91 samples like this: 80 L. panamensis, 11 L. braziliensis and 7 indeterminate, these indeterminate samples were sent for sequencing and it was established that 4 of them are more associated with L. panamensis sequences and 3 of them show an intermediate relationship between L.. panamensis and L. braziliensis.

**Image 2.**
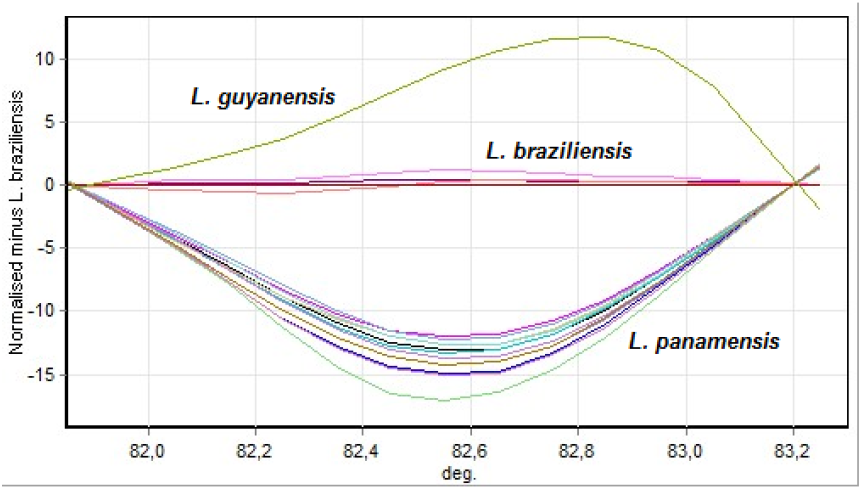
Analysis of PCR-HRM data using differential graphs, using DNA from L. braziliensis, L. panamensis and L guyanensis as reference.

### Reproducibility of HRM PCR typing

Each of the samples was run in triplicate in 3 different runs and no discordances were observed in the typing.

## 4. DISCUSSION

*Leishmaniasis* is a parasitic disease with worldwide distribution.(1). Colombia is the second country with the highest number of cases in Latin America after Brazil(11).

Diagnosis in Colombia and in most countries in the world is necessary to start treatment because the medications used are highly toxic and are contraindicated in pregnant women, children with malnutrition, and people with liver or kidney disorders.(12).

The diagnostic methods indicated by the WHO are: direct examination, which has variable sensitivity (50-90%) and is influenced by different factors such as quality and type of sample and the experience of the person doing the reading.(13), the culture has a much lower sensitivity (10-30%) and the growth of the parasite can take up to a month, which prevents prompt implementation of the treatment. Furthermore, the chance of being able to isolate the parasite in culture is inversely proportional to the time of treatment. evolution of the disease, because perhaps the parasite load is reduced(14).

In view of the need to implement rapid, sensitive and specific methodologies, progress has been made in molecular diagnosis, based mainly on the detection of parasite DNA, it has emerged as a rapid and reliable diagnostic alternative with high sensitivity and specificity.(15).

However, molecular diagnosis is one thing and species identification is another. This identification is basically necessary for 3 reasons: 1 understanding the epidemiology and distribution of the disease, 2 identification of resistant strains, 3 choosing the appropriate treatment.

The identification of species allows: to characterize parasite populations to improve diagnosis and treatment, evaluate relapses or reinfections, and establish prognosis and control.

Although one of the main advances in identification using PCR RFLP(16), amplifying a region of (593 bp) amplifying the Hsp70 gene, with which it has been possible to differentiate 9 species using a single pair of oligonucleotides and a single HaeIII restriction enzyme, however when trying to resolve the cutting patterns in some cases By presenting differences of only 10 nucleotides in the banding pattern between one species and another, differentiation in the agarose gel is difficult.

Advances in species identification using PCR RFLP have been based on the presence of a large number of single nucleotide polymorphisms in the genome of *Leishmania* spp.(17).

Taking into account the presence of these polymorphisms and the ability to identify these differences using PCR HRM, which first of all is a much more sensitive technique, since in our case it managed to detect infection at low parasite loads as much as 1 fg of parasite DNA, while conventional PCR is only achieved with 1000 fg of the parasite’s DNA, achieving a vast improvement in analytical sensitivity.

The DNA polymerase 2 gene was chosen because it is a single-copy gene and since it is a single-copy gene, no differences in the sequence will be observed in the same parasite, as occurs in the multiple-copy genes that have been used. in the typing(18).

The in silico analysis of a region of the DNA polymerase 2 gene allowed us to determine differences at the nucleotide level that may occur between species and according to the properties of PCR-HRM to differentiate change in fluorescence based on temperature curves. High resolution medium denaturation allowed the final realization of species differentiation(19).

The PCR-HRM made it possible to simultaneously detect and identify the 3 species of *Leishmania* responsible for more than 98% of the cases of cutaneous *Leishmaniasis* in Colombia. Additionally, the typing by this technique does not present discrepancies with those obtained by PCR-RFLP and the results are reproducible.

The specificity test showed the absence of nonspecific amplification products and oligonucleotide dimers as observed by the melting curves, in addition to an amplicon size consistent with that expected, demonstrating that they amplified the region for which they were designed and that they did not present alignments. nonspecific.

Furthermore, with these oligonucleotides and by normalizing the fluorescence, high-resolution average denaturation temperature curves were obtained that were specific for each species evaluated, thus allowing their correct identification and subsequent analysis with the respective clinical samples.

The analytical sensitivity of both PCRs (HSP70N PCR and PCR-HRM) reaffirmed what was described by previous studies.(15)in which a greater sensitivity of real-time PCR was demonstrated compared to conventional PCR, most likely due to the detection method used in conventional PCR.

Evaluating the linearity of the PCR-HRM, it was observed that it presented good linearity in the different concentrations analyzed, which allows us to affirm that the technique has a directly proportional relationship between the concentration of the parasite DNA and the amplification cycle. This added to the correlation coefficient and high efficiency which had already been described in previous studies for PCR-HRM.(15). For its part, the reproducibility test did not show changes in the diagnoses and/or classifications, evidencing the reliability of the results. This is thanks to the technique’s ability to eliminate intra-assay variations, concentrating the analyzes on the common points between the samples and the controls.

The evaluation of the interference of human DNA in the amplification efficiency revealed a slight decrease in the efficiency of real-time PCR in those samples with low levels of parasite DNA. However, amplification was obtained in a greater number of samples compared to conventional PCR and that were parasitologically positive, according to direct examination, without observing changes in the typing. This allowed obtaining a positive diagnosis of all samples despite the presence of human DNA, which is known to affect DNA amplification in PCR reactions.(20).

In this case, the sensitivity of the direct diagnosis was 100% because the personnel who performed the direct diagnosis are highly trained. The sensitivity shown for the diagnosis with HRM PCR was 100%, with a specificity of 100%, there were no false negatives or false positives, which shows the excellent performance of this technique as a diagnostic test.

By evidencing the advantages that real-time PCR has over other diagnostic methods(20,21), and complemented with high-resolution melting curves for species identification, there is initially an optimization of the time to obtain a diagnosis and identification in addition to reducing the percentage of indeterminate samples from 16% to 7.14%.

It is the first work in Colombia that uses PCR HRM for the identification of *Leishmania*. Additionally, to date, there are few reports in the world using this PCR-HRM methodology for the identification of *Leishmania* in patients.(3,18–21). The implementation of this work will allow its application in diagnosis, clinic and epidemiology. The applications are aimed at diagnostic laboratories, reference and research centers, which require reliable tools for the detection and identification of *Leishmania* spp. This proposal is aimed at contributing to the development and implementation of new diagnostic strategies that can favor the implementation or even to monitor the effectiveness of measures for the prevention, treatment and control of *Leishmaniasis* and achieve a decrease in the incidence of this disease, being able to adapt for the typing of strains responsible for other clinical forms of *Leishmaniasis* such as mucosal, diffuse, disseminated and visceral.

## Data Availability

All data produced in the present study are available from the authors upon reasonable request.
All data produced in the present work are contained in the manuscript.

